# Modelling COVID-19 Transmission in the United States through Interstate and Foreign Travels and Evaluating Impact of Governmental Public Health Interventions

**DOI:** 10.1101/2020.05.23.20110999

**Authors:** Nita H. Shah, Nisha Sheoran, Ekta Jayswal, Dhairya Shukla, Nehal Shukla, Jagdish Shukla, Yash Shah

**Author notes:** Email: ^1^, ^2^, ^3^ ^4^, ^5^, ^6^, ^7^. Corresponding Author: ^6^ Jagdish Shukla, MD.

## Abstract

**Background:** The first case of COVID-19 was reported in Wuhan, China in December 2019. The disease has spread to 210 countries and has been labelled as pandemic by WHO. Modelling, evaluating, and predicting the rate of disease transmission is crucial for epidemic prevention and control. Our aim is to assess the impact of interstate and foreign travel and public health interventions implemented by the United States government in response to the Covid-19 pandemic.

**Methods:** A disjoint mutually exclusive compartmental model is developed to study transmission dynamics of the novel coronavirus. A system of non-linear differential equations was formulated and the basic reproduction number R_0_ was computed. Stability of the model was evaluated at the equilibrium points. Optimal controls were applied in the form of travel restrictions and quarantine. Numerical simulations were conducted.

**Results:** Analysis shows that the model is locally asymptomatically stable, at endemic and foreigners free equilibrium points. Without any mitigation measures, infectivity and subsequent hospitalization of the population increases while placing interstates individuals and foreigners under quarantine, decreases the chances of exposure and subsequent infection, leading to an increase in the recovery rate.

**Conclusion:** Interstate and foreign travel restrictions, in addition to quarantine, help in effectively controlling the epidemic.

## 1. Introduction

In December 2019, an unidentified pneumonia was found in Wuhan, Hubei province, China. The responsible virus was later identified as the novel coronavirus 2019, and the disease as coronavirus disease 2019 (COVID-19) (Cohen and Normile 2020). COVID-19 is transmitted via direct contact with an infected person through respiratory droplets when a person coughs or sneezes, or by indirect contact with contaminated surfaces with respiratory droplets from infected person and then touching their eyes, nose or mouth(Holshue et al. 2020). Furthermore, the virus remains on surfaces from few hours to several days and has an incubation period between 1-14 days. Consequently, the disease spread rapidly from Wuhan to all parts of the country and overseas.

Introduction and spread of Coivd-19 within the United States is a direct result of transmission through foreign and interstate travel. The first known case of COVID-19 in U.S.A. was confirmed on 20^th^ January 2020 in a 35-year-old individual who had travelled from Wuhan to Washington state(Holshue et al. 2020). Soon, cases started appearing and rising in many other states of U.S.A. including New York, New Jersey, Illinois, Florida, Georgia, Texas, Pennsylvania due to interstate travel. The CDC alarmed that hospital may get overwhelmed by large number of people seeking care at the same time due to widespread transmission of disease which may lead to otherwise preventable deaths (2020). In response to the oncoming epidemic the US government implemented the following regulations:

**Table 1:** Timeline of Public Health Intervention Implemented by the United States Government.

| **Date** | **Action** |
| --- | --- |
| 17<sup>th</sup> Jan | Public health entry screening at 3 U.S. Airport |
| 31<sup>st</sup> Jan | Corona Virus Public health emergency, Chinese travel restrictions, restricted entry into U.S.A for foreign nationals who poses risk of transmission, Funnel all flight from China to just 7 U.S. domestic airport. |
| 29<sup>th</sup> Feb | Barred all travel to Iran, level 4 travel advisory to areas of Italy and South Korea. |
| 11<sup>th</sup> March | Travel restriction for foreigners who visited Europe in last 14 days. |
| 14<sup>th</sup> March | Europe travel ban extended to UK and Ireland. Implement Social Distancing and Closure of teaching institutes in many states. |
| 18<sup>th</sup> March | Temporary closure of U.S. Canada border for non- essential traffic |
| 19<sup>th</sup> March | Americans to avoid all international travel |
| 20<sup>th</sup> March | U.S. and Mexico agreed to restrict non-essential cross border traffic.<br>Closure of non-essential businesses and shelter in place order in NY. |
| 24<sup>th</sup> March | Self-quarantine for 14 days for individual who recently visited New York. |
| 28<sup>th</sup> March | For residents in NY, NJ, CT, Avoid non-essential domestic travel for 2 weeks. |
| 29<sup>th</sup> March | Social distancing extended through 30<sup>th</sup> April. |
| 3<sup>rd</sup> April | All American wear non-medical, fabric or cloth mask to prevent asymptomatic spread of coronavirus. |

While the United States has implemented numerous public health interventions, it has not implemented a ban on interstate travel. According to World Health Organization (WHO), New cases of COVID-19 have emerged in 210 countries with 1,733, 945 confirmed cases and 106, 518 confirmed deaths globally as of 10^th^ April 2020. The United States has implemented quarantine measures, close contact tracing, early testing for individuals with symptoms, hospitalization if needed, and closing of teaching institutes and non-essential businesses. Studies have shown that in other countries, the complete lockdown of travel has decreased the spread of the disease in the surrounding states (Castillo-Chavez et al. 2003; Tang et al. 2020). In order to prevent the transmission of Covid-19 within the US, the mode of transmission must first be modeled and understood. Mathematical modeling is ideal for evaluating and predicting the rate of disease transmission. Data-driven mathematical modelling plays an important role in epidemic mitigation, in preparedness for future epidemic and in the evaluation of control effectiveness. (Wu et al. 2020). In this study, we adopted a disjoint mutually exclusive compartmental model to shed light on the transmission dynamics from foreign and interstate travel of the novel coronavirus and our aim is to assess the impact of public health interventions on infection by measuring basic reproduction number, contact rate, newly confirmed cases, total confirmed cases, total death. Our estimated parameters are largely in line with World Health Organization estimates and previous studies (2019).

## 2. Mathematical Modeling

Mathematical modeling plays vital role in determining dynamics of diseases. In this paper we consider disjoint mutually exclusive compartmental model with compartments as follows:

Exposed to Covid-19 *E*, Covid-19 transmission in interstate *I_S_*, Covid-19 transmission through foreigners *F*, Quarantined class *Q*, Covid-19 Infected *I*, Hospitalized *H*, and Recover *R*.

Notations and parametric values used in the formulation of dynamical system model are given in the following Table 2.

**Table 2:** Parametric definitions and its values

| Notations | Description | Parametric values |
| --- | --- | --- |
| $B$ | Birth rate | 0.0181 |
| $\beta_1$ | Rate at which interstate population gets exposed to Covid-19 | 0.0011 |
| $\beta_2$ | Rate at which foreigner gets exposed to Covid-19 | 0.0003 |
| $\beta_3$ | Rate at which Foreigner comes in contact with interstate population | 0.0012 |
| $\beta_4$ | Rate at which Interstate population goes for quarantine | 0.0018 |
| $\beta_5$ | Rate at interstate population gets infected | 0.0035 |
| $\beta_6$ | Rate at which foreigner quarantine themselves | 0.000015 |
| $\beta_7$ | Rate at which foreigner gets infected | 0.000001 |
| $\beta_8$ | Rate at which quarantine humans gets infected by Covid-19 | 0.0025 |
| $\beta_9$ | Rate at which infected humans gets hospitalized | 0.0037 |
| $\beta_{10}$ | Rate at which exposed humans gets hospitalized | 0.000002 |
| $\beta_{11}$ | Rate at which hospitalized humans gets recovered | 0.0000003 |
| $\mu$ | Death rate | 0.000119 |
| $\mu_c$ | Death rate due to Covid-19 | 0.0027 |

This model considers new recruitment in the exposed class at the rate *B* and all the compartments have mortality rate *μ*. Here interstate population (it is the population within the country) is exposed to Covid-19 at the rate *β*_1_ and Foreigner population (it is population arriving in a particular country through air ways) is exposed to Covid-19 at the rate *β*_2_. Next, foreigners *F* come in contact with interstate population at the rate *β*_3_ joining *I*_S_. Interstate population and foreigners quarantine themselves at the rate *β*_4_ and *β*_6_ respectively. Similarly, after getting exposed to Covid-19, interstate and foreigner population gets the infection joining infectious class *I* with the rate *β*_5_ and *β*_7_ respectively. Quarantined humans also get the infection at the rate *β*_8_. Next, we assume infected population gets hospitalized at the rate *β*_9_ joining *H*. We also assume population gets admitted to the hospital at the initial exposure of the disease at the rate *β*_10_. Hospitalized patients after undergoing treatment gets recover joining *R* at the rate *β*_11_ We take into the consideration death due to Covid-19 when the individual is hospitalized.

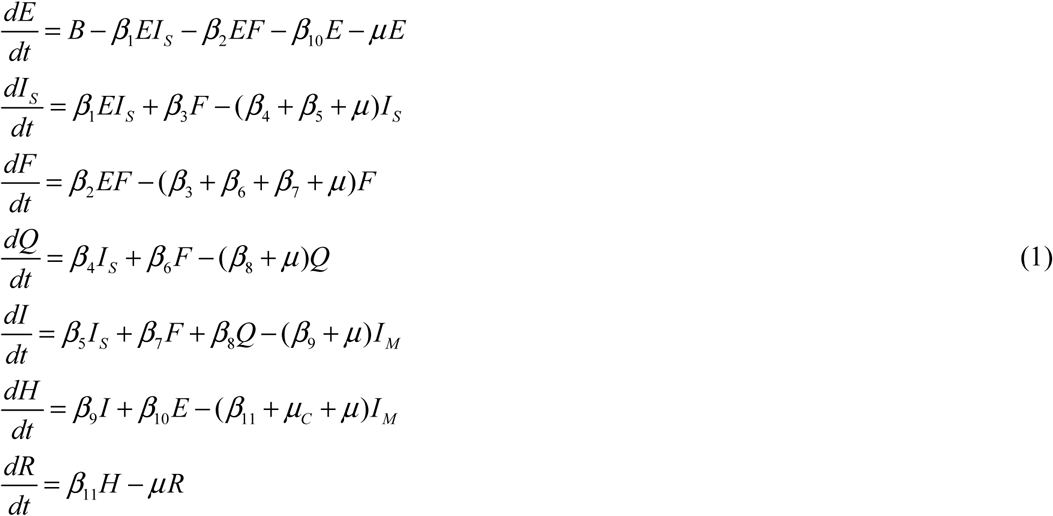

where, *N*(*t*) = *E*(*t*) + *I_S_* (*t*) + *F* (*t*) + *Q*(*t*) + *I* (*t*) + *H* (*t*) + *R*(*t*)

The Fig. 1 gives rise to the following set of non-linear ordinary differential equations

**Fig. 1.**
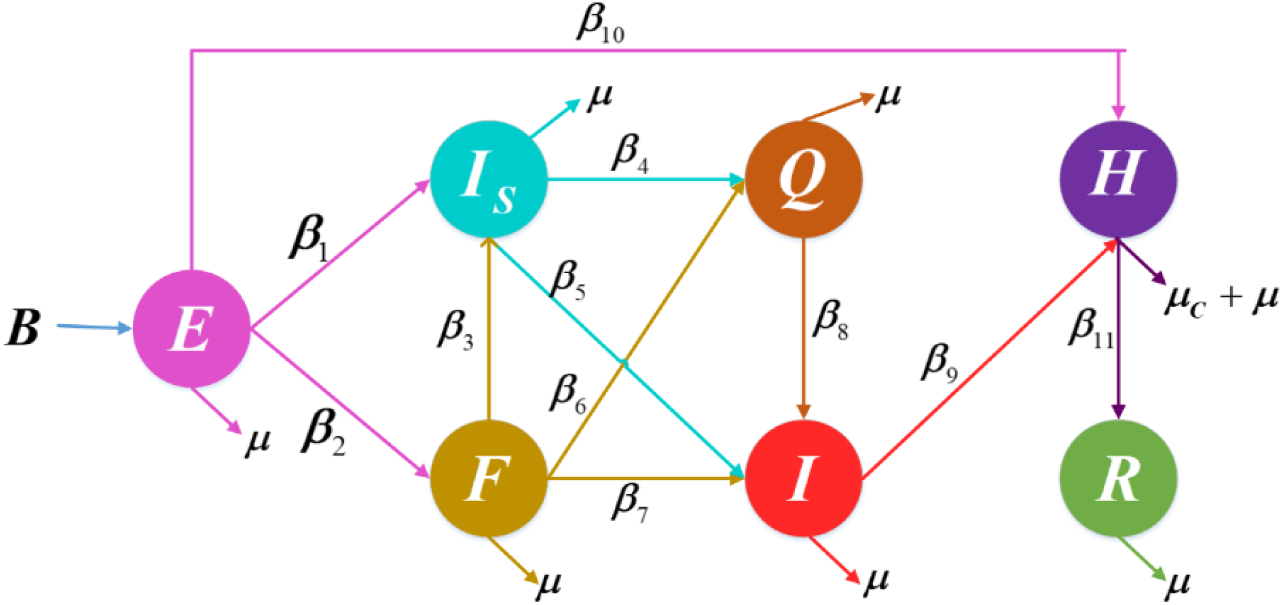
Movement of individuals from one compartment to another compartment

Adding all the differential equations of model (1), we get,

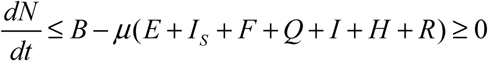

Hence, 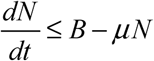, So that 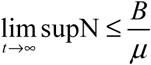.

Then, feasible region for the system is defined as

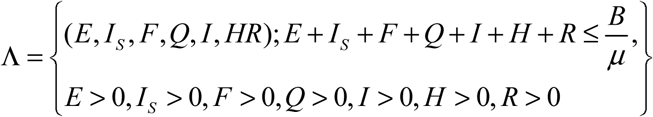

This system has following equilibrium points

i. Foreigner free equilibrium point
ii. Endemic equilibrium point

## 3. Reproduction number

Basic reproduction number is defined as the total number of secondary infections in a total susceptible population.

Here, we calculate reproduction number using Diekmann *et al*, when the disease in its endemic stage i.e. for this model it is defined as percentage of population infected by a single infection in a totally exposure situation(Diekmann et al. 2010).

We also compute the value of reproduction number *R_F_* when there are no foreigners present in the total population.

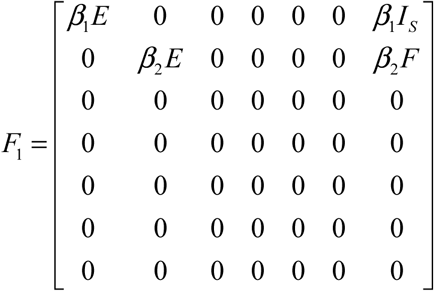

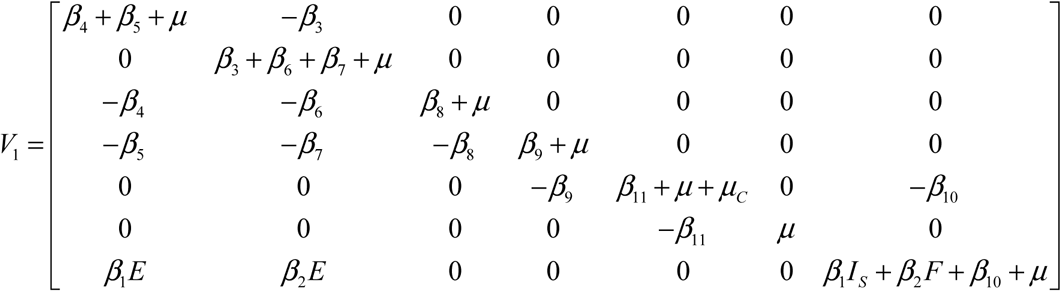

The reproduction number 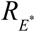, 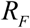 is the spectral radius of 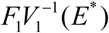 and 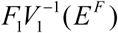 respectively. The value of 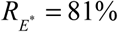 and *R_F_* = 1.10.

## 4. Stability analysis

In this section we study the stability analysis of the model. Here we study Local stability of all the equilibrium points using Routh-Hurwitz criterion by Routh 1877 (Routh).

### Theorem 1

The foreigner free equilibrium point is locally asymptotically stable if

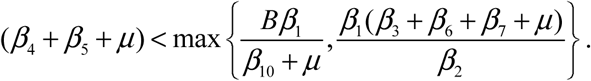

#### Proof

The Jacobian of system (1) at Foreigner free equilibrium is as follows

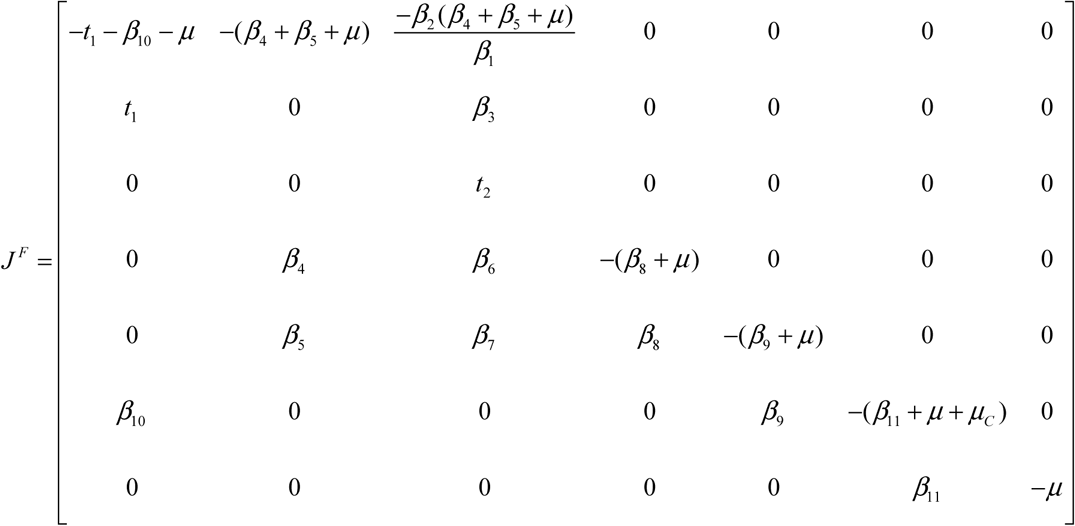

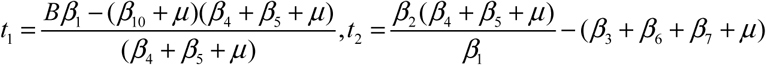

The eigen values of the Jacobian *J^F^* are

*λ*_1_ = *t*_2_, *λ*_2_ = −(*β*_8_ + *μ*), *λ*_3_ = −(*β*_9_ + *μ*), *λ*_4_ = −*μ*, *λ*_5_ = −(*β*_11_ + *μ* + *μ_C_*), 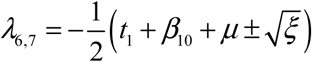, 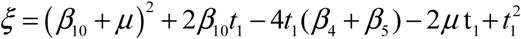

If it has imaginary roots i.e. *ξ* < 0, Then we have negative real part. Hence the theorem. But if *ξ* ≥ 0, then eigen values are negative if

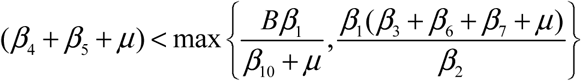. Hence the Foreigner free equilibrium point is locally asymptotically stable.

### Theorem 2

The endemic equilibrium point is locally asymptotically stable if 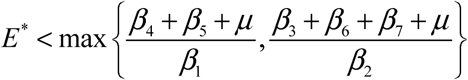

#### Proof

The Jacobian matrix of system (1) for endemic equilibrium is given by

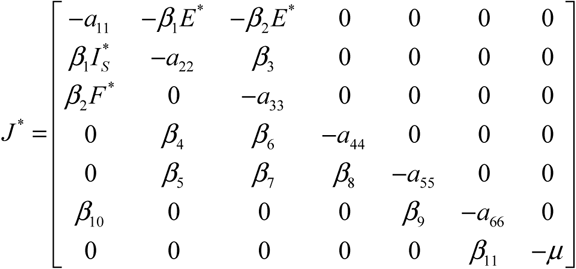

where,

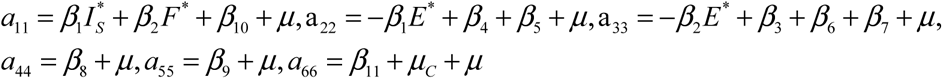

The characteristic polynomial for Jacobian *J*^*^ is

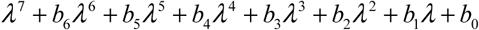

where,

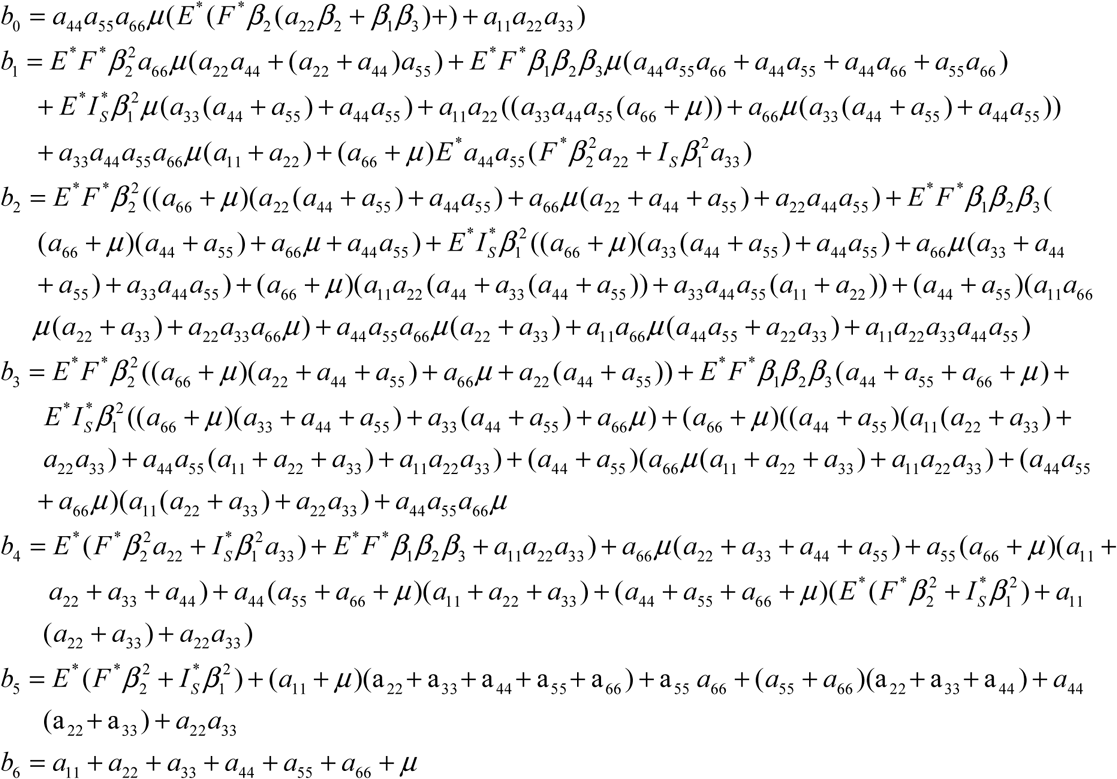

Here, all the eigen values are negative if a_11_ > 0, a_22_ > 0, a_33_ > 0, a_44_ > 0, a_55_ > 0, a_66_ > 0 i.e.

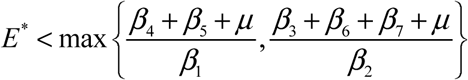. Then by Routh-Hurwitz criterion we say the endemic equilibrium point is locally asymptotically stable.

## 5. Optimal control

The novel corona virus is spread through human contact with infected individuals. Therefore, one can put control on respective situation to prevent its spreading.

Control description:

*u*_1_: To prevent exposed foreign individuals in the interstate
*u*_2_: Exposed interstate individuals should be quarantined
*u*_3_: Exposed foreign individuals should be quarantined
*u*_4_: Infected individuals should be quarantined

The objective function is,

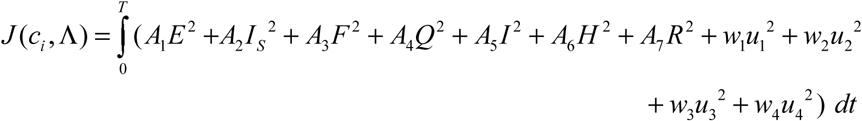

where, Λ denotes set of all compartmental variables, *A*_l_, *A*_2_, *A*_3_, *A*_4_, *A*_5_, *A*_6_, *A*_7_ denote non-negative weight constants for compartments *E, I*_S_, *F*, *Q, I, H, R* respectively. *w*_1_, *w*_2_, *w*_3_ and *w*_4_ are the weight constants for each control *u_i_* where *i* = 1,2,3,4 respectively.

Now, calculate every values of control variables from *t* = 0 to *t = T* such that,

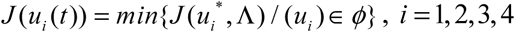

where, *ϕ* is a smooth function on the interval [0,1].

Related Langrangian function is given by,

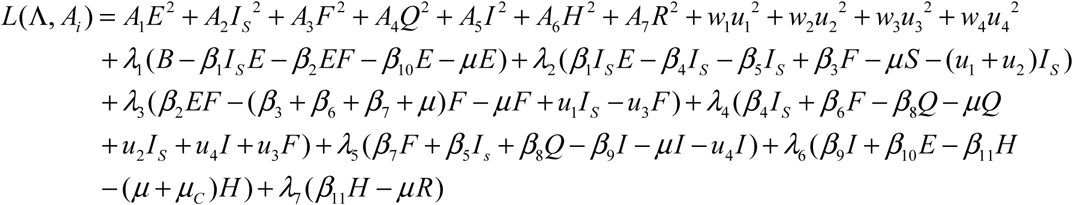

The adjoint equation variables, *λ_i_* = (*λ*_1_, *λ*_2_, *λ*_3_, *λ*_4_, *λ*_5_, *λ*_6_, *λ*_7_) for the system is calculated by taking partial derivatives of the Langrangian function with respect to each compartment variable.

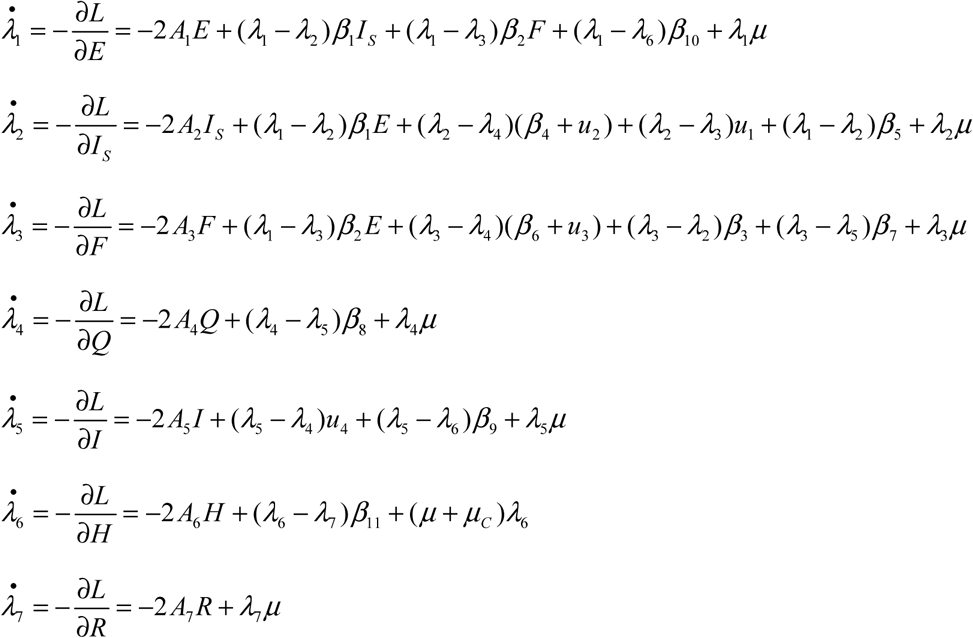

This calculation leads with resulting conditions as (Pontryagin 2018),

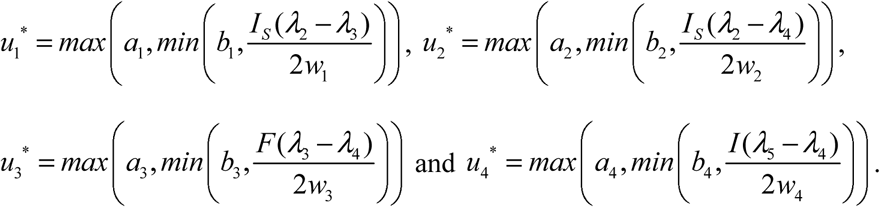

Based on analytical results, numerical simulation is given in next section.

## 6. Numerical simulation

In this section we discuss the simulation performed for the system (1).

From fig.2. we observe 30% of interstate population is exposed to Covid-19 in 17.2 days. Whereas 24.55% of foreigner’s population is exposed to Covid-19 in 21.8 days. 21% of foreigners come in contact with Interstate individuals in 7.1 days which increases the infectives of interstate to 22.78% in 9.2 days. Also 27.59% of interstate population gets hospitalized in 14.5 days.

**Fig. 2.**
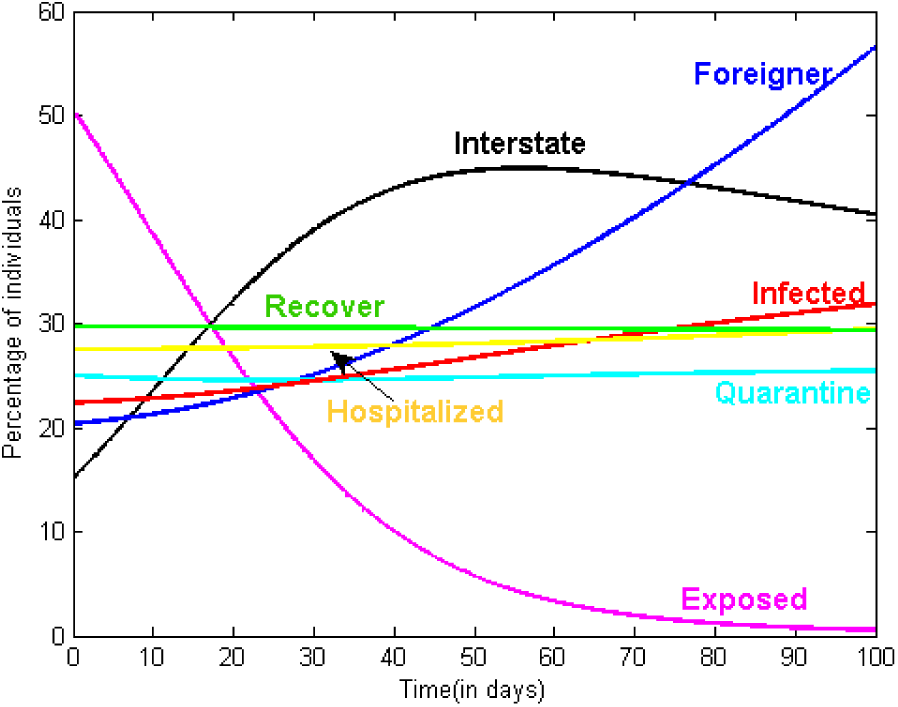
Transmission of individuals in various compartments

Scatter plotting is shown in figure 3. Combined effect of group of three compartments is revealed in each plot. Fig. 3(a) depicts that; more infected interstate and foreign individuals will be hospitalised at higher rate of level. Fig. 3(b) shows that, individuals who travelled more will be quarantined. From figure 3(c), one can say that infected foreigner would be quarantined at higher rate. All quarantined individuals infected more than, they get hospitalised which is observed in figure 3(d). Figure 3(e) describes that how interstate infected individuals are quarantined.

**Fig. 3.**
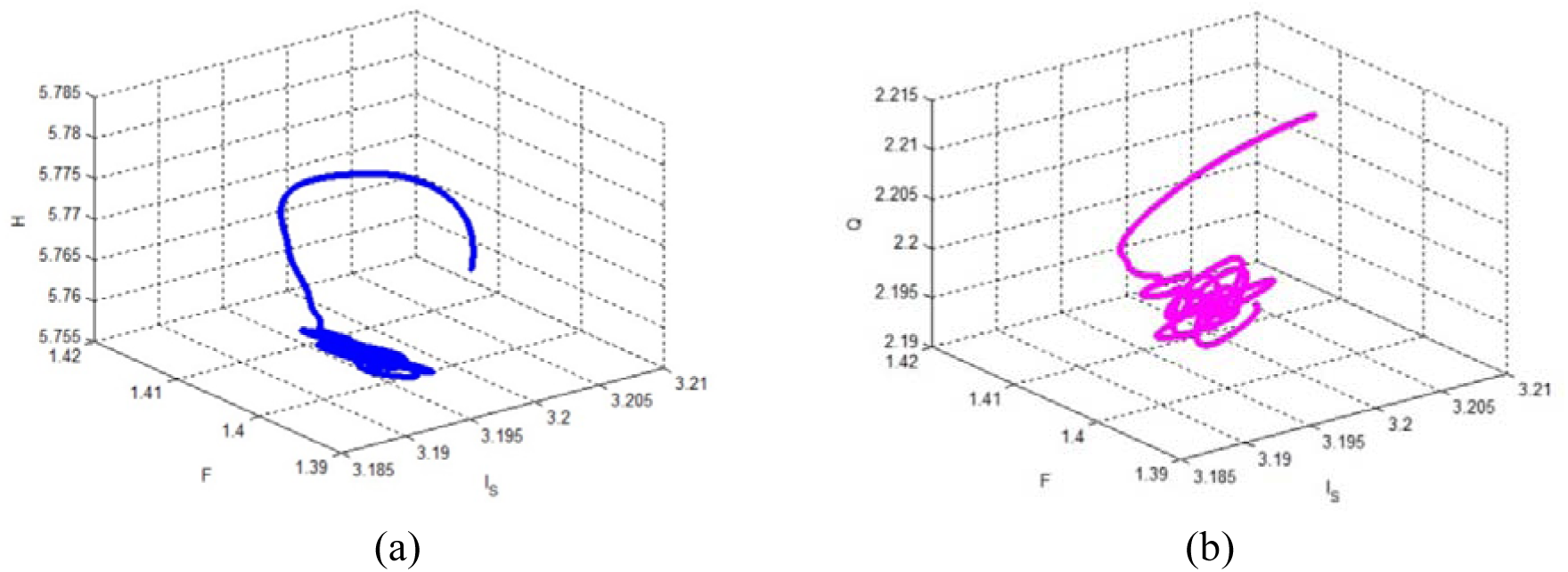
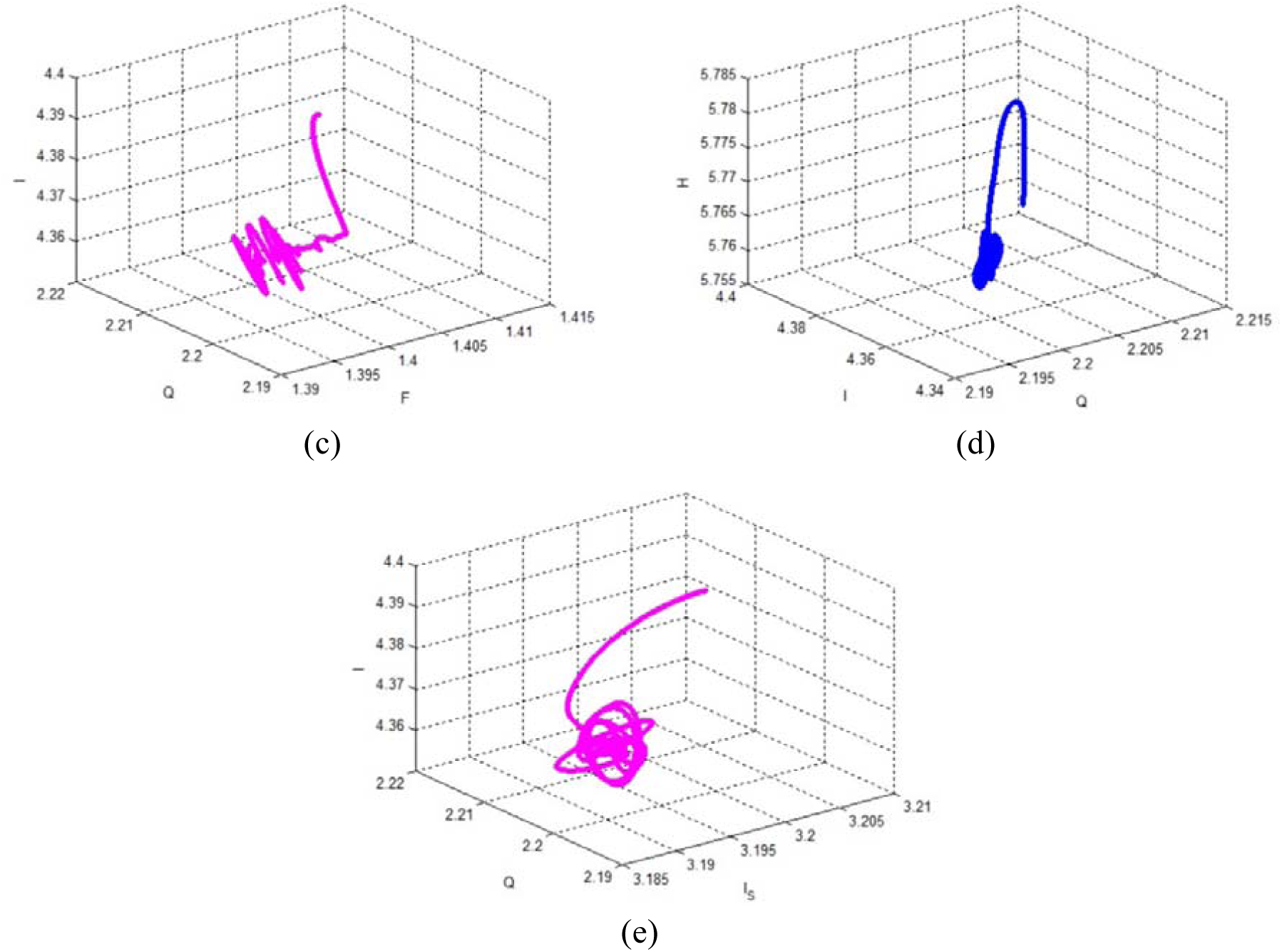
Scatter plotting

Fig.4 shows the periodic nature of the interstate class exposed to the virus Covid-19. It indicates that interstate population is exposed again and again to the disease. It happens if the lockdown, social distancing is not followed as per government system. Which shows the importance of the government action taken against Covid-19 to protect the population. Fig.5 Shows the stability of the respective compartments at endemic equilibrium point. Since the government has decided to quarantine foreigners as soon as they arrive in their countries this makes the system stable as they are not exposed much to the Covid-19.

**Fig. 4.**
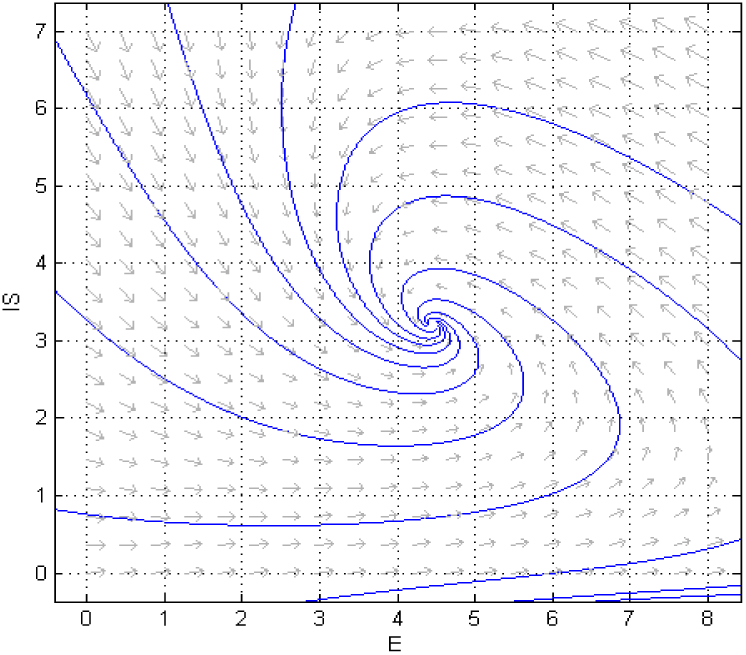
Phase plot of interstate class when exposed to Covid-19

**Fig. 5.**
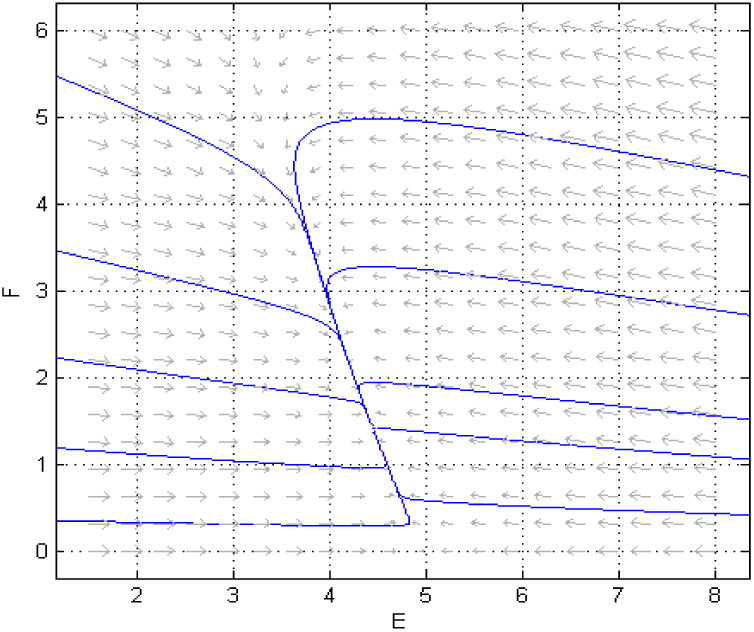
Phase plot of foreigner when exposed to covid-19

Figs. 6-1, 6-2 shows the trajectory at the endemic equilibrium point for the system (1). Here, we observe the importance of quarantine as the system is stable when interstate and foreigners are quarantined.

**Fig. 6-1.**
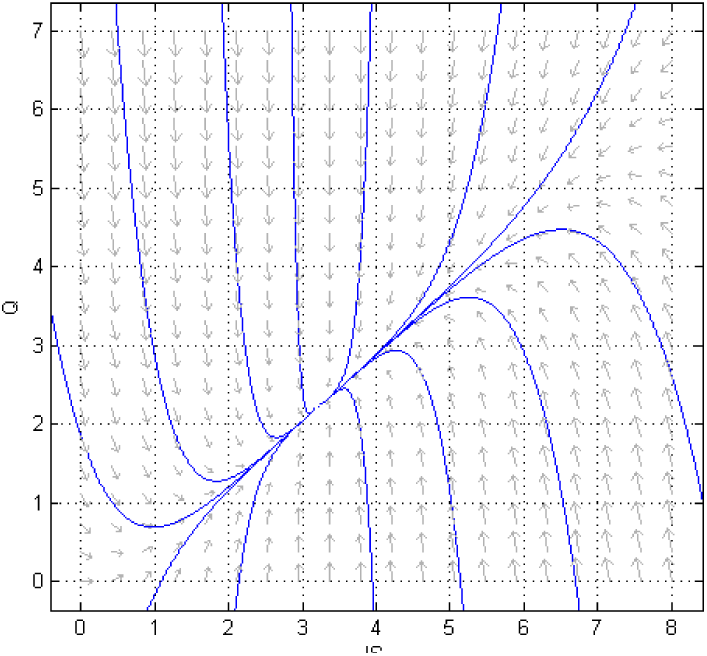
Behaviour of interstate class with quarantine class

**Fig. 6-2.**
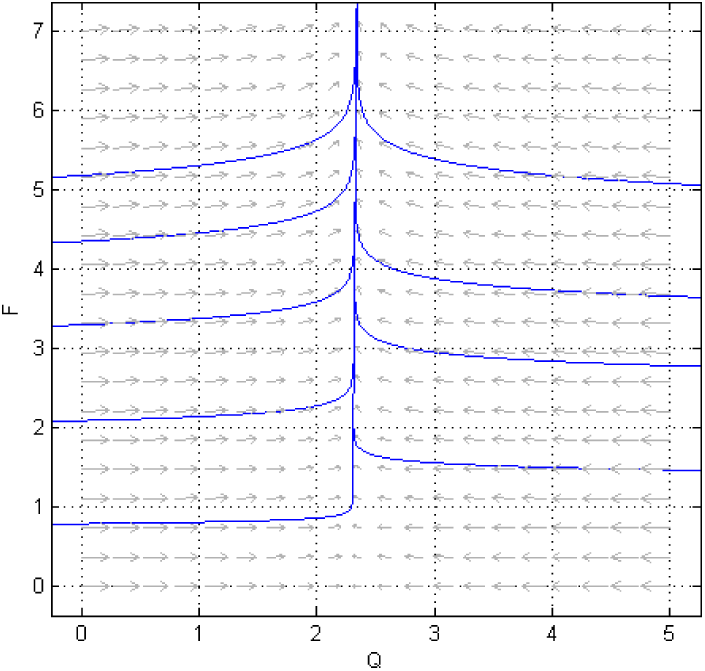
Phase plot of quaratine with foreingners

From fig.7, we observe foreigner are moving towards interstate population.

**Fig. 7.**
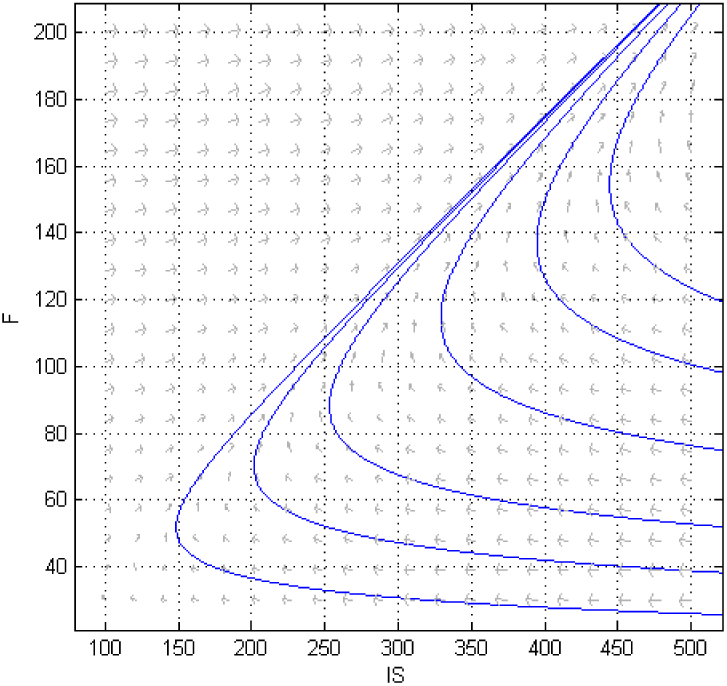
Transition diagram of interstate and foreigner population

Fig.8-9 illustrates the flow of interstate population and foreigners with the infection. It shows that the interstate population gets the infection slowly as compared to foreigners.

**Fig. 8.**
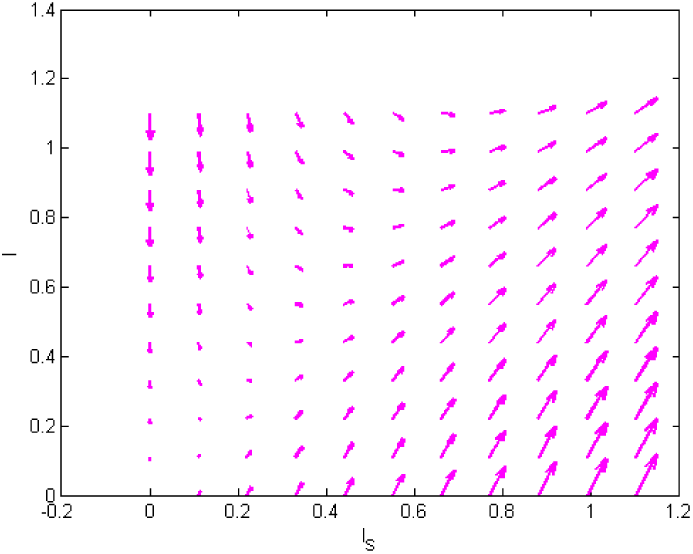
Directional plot of Interstate population infected with Covid-19

**Fig. 9.**
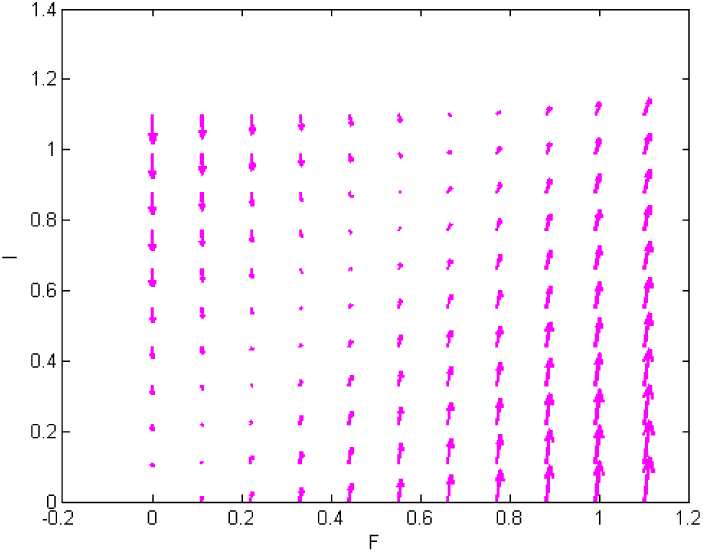
Behaviour of Foreigner population infected with Covid-19

Fig.10(a), 10(b) show the flow of interstate and foreigners towards hospitalization. Foreigners gets hospitalized at faster rate than interstate population.

**Fig. 10.**
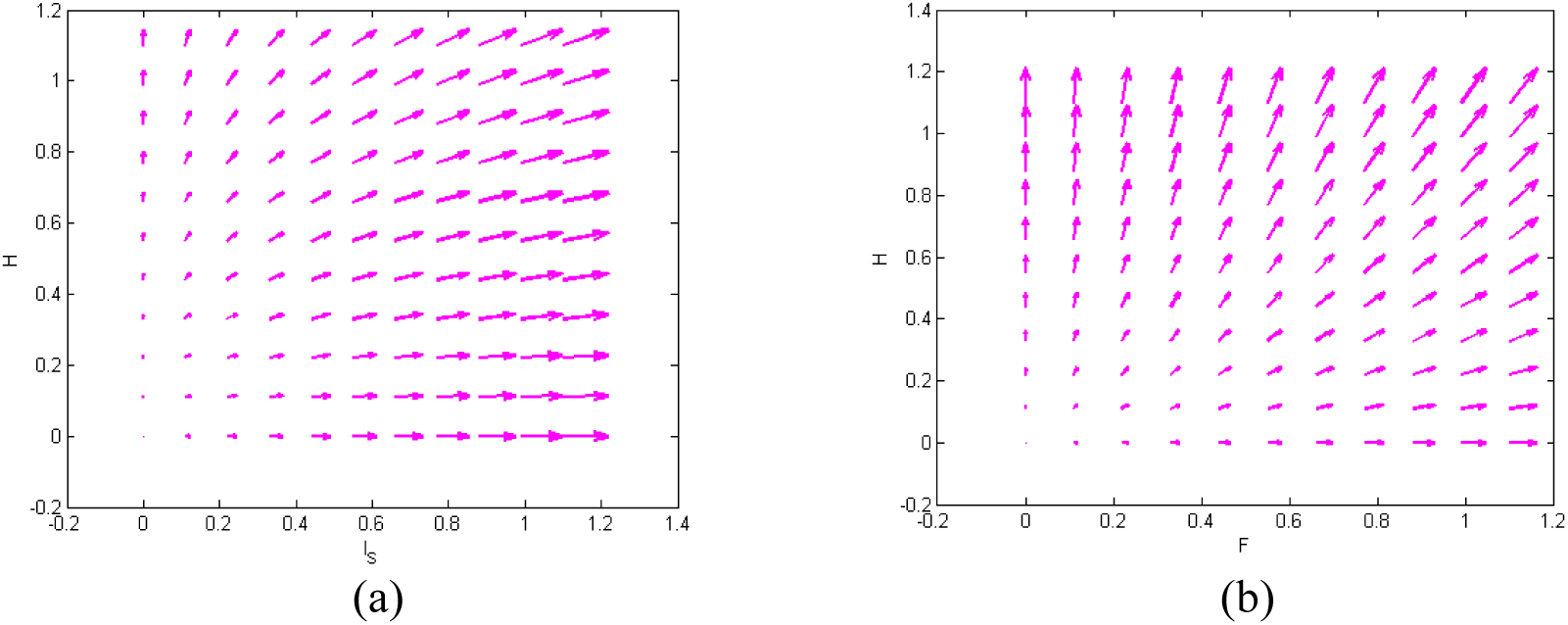
Directional plot of hospitalization of interstate population and foreigner

Figures 11 (a)-(g) show the oscillating behaviour of each compartment. As epidemic nature of disease increases, this can oscillate whole situation of model. In some interval of data, exposed individuals increase (Fig. 11(a)) who are either interstate (Fig. 11(b)) or foreigner (Fig. 11(c)). If quarantined individuals do not follow quarantines rules which has been observed in Fig. 11(d). This leads to a greater number of infected individuals (Fig. 11(e)) hence they should be hospitalised (Fig. 11 (f)) which effects on recovery rate (Fig. 11(f)).

**Fig. 11.**
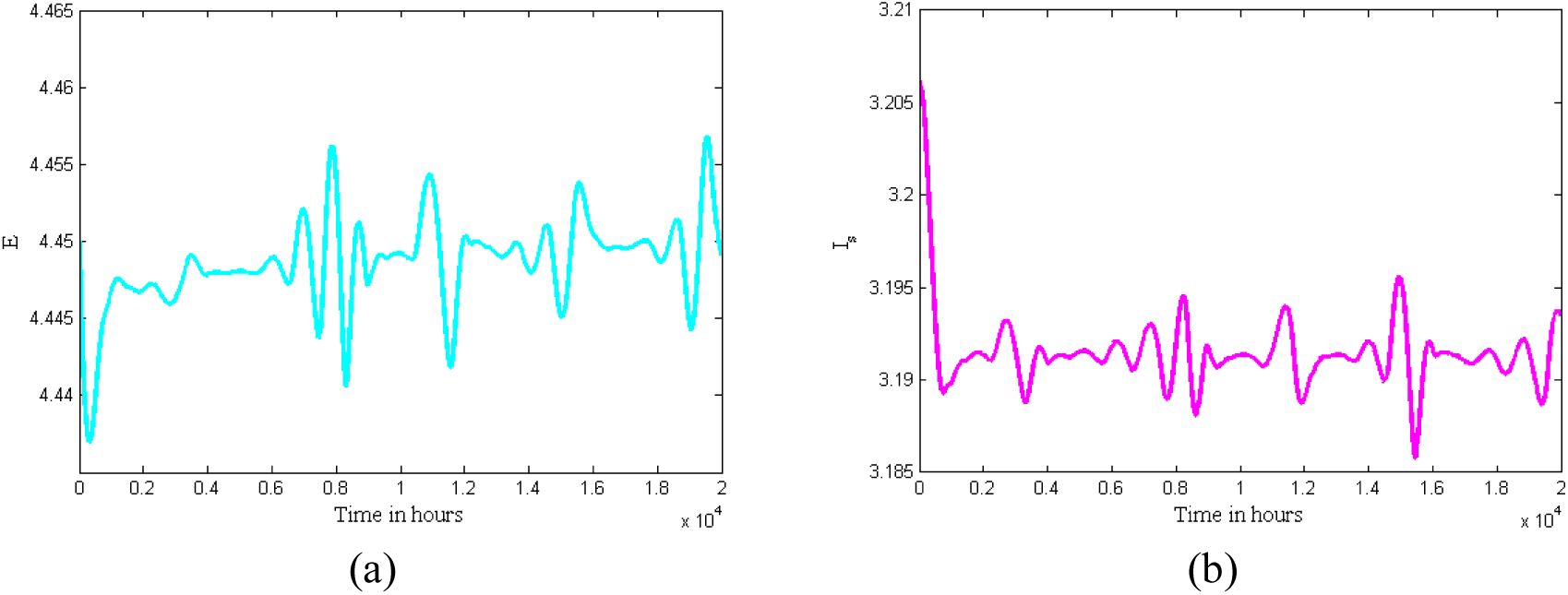
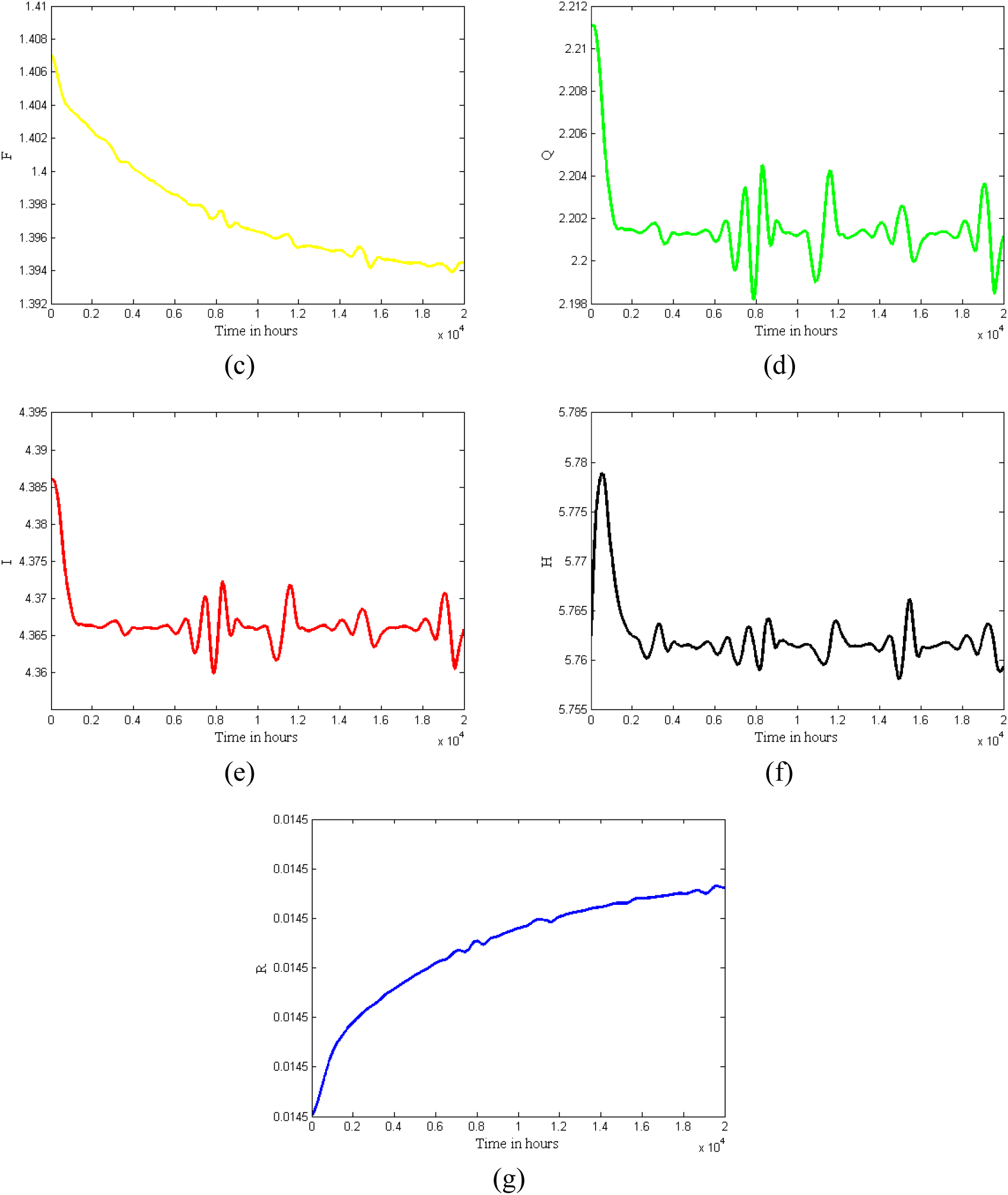
Oscillations Effect

The above oscillating nature of the model is controlled by the fig. 12 (a)-(g). All the four controls are effective to this model. Control applied decreases the number of exposed individuals. Quarantining interstate and foreign individuals reduce the infection leading to increase in recovery rate.

**Fig. 12.**
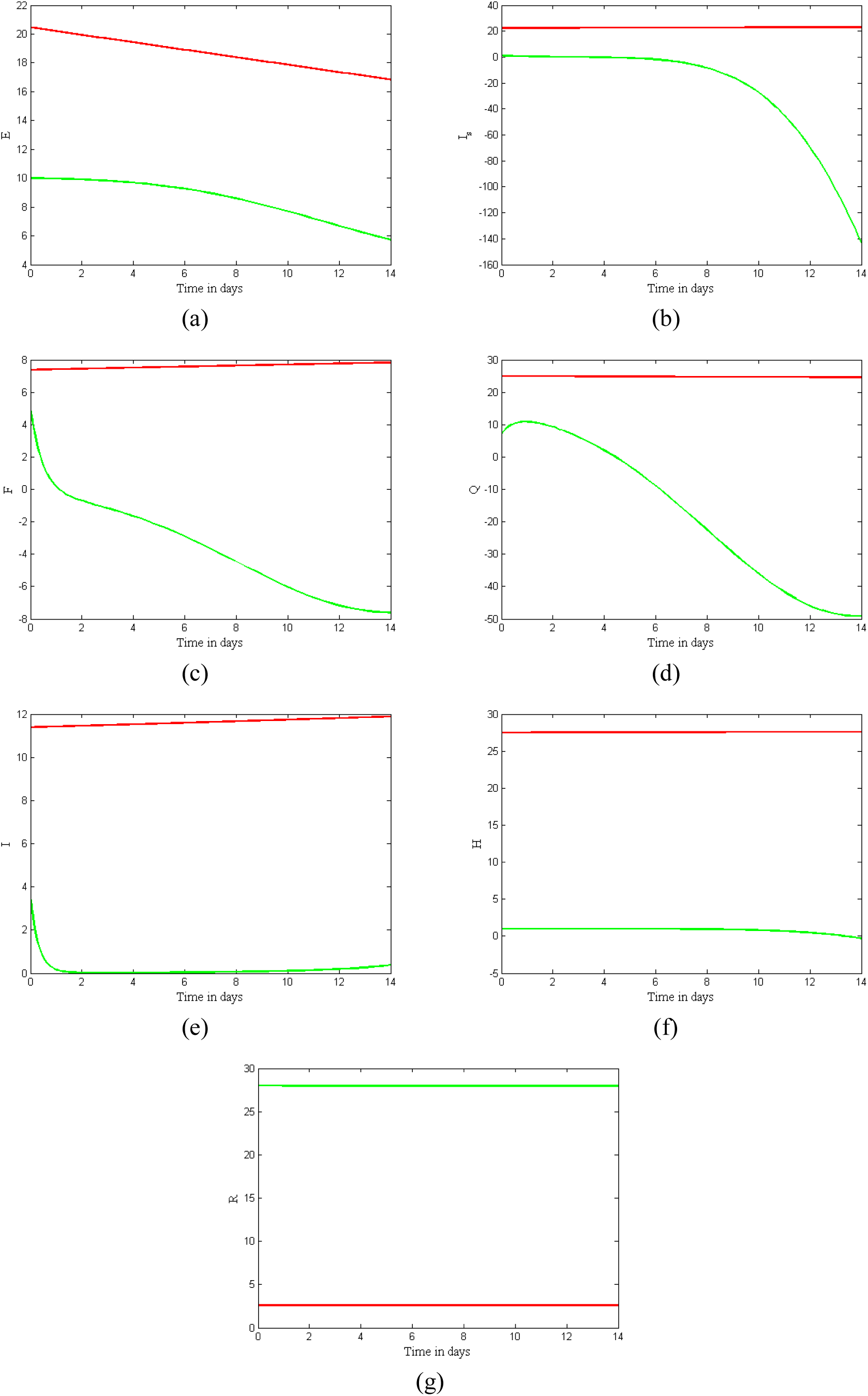
Control Effect on the Model

The above oscillating nature of the model is controlled by the fig.12(a)-(g). All the four controls are effective to this model. Control applied decreases the number of exposed individuals. Quarantining interstate and foreign individuals reduce the infection leading to increase in recovery rate.

**Fig. 13.**
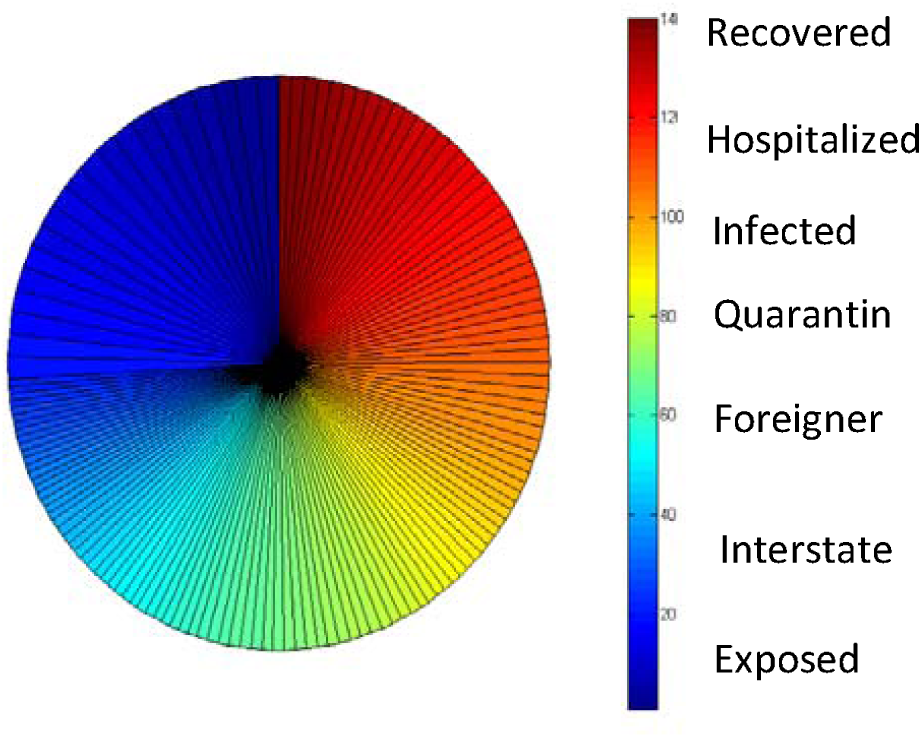
Chaotic Diagram showing mortality rate of 2019-nCoV

The fig.14(a)-(b) clearly shows the infected population of foreigner is more than that of interstate population which show the importance of complete ban on air arrivals.

**Fig. 14.**
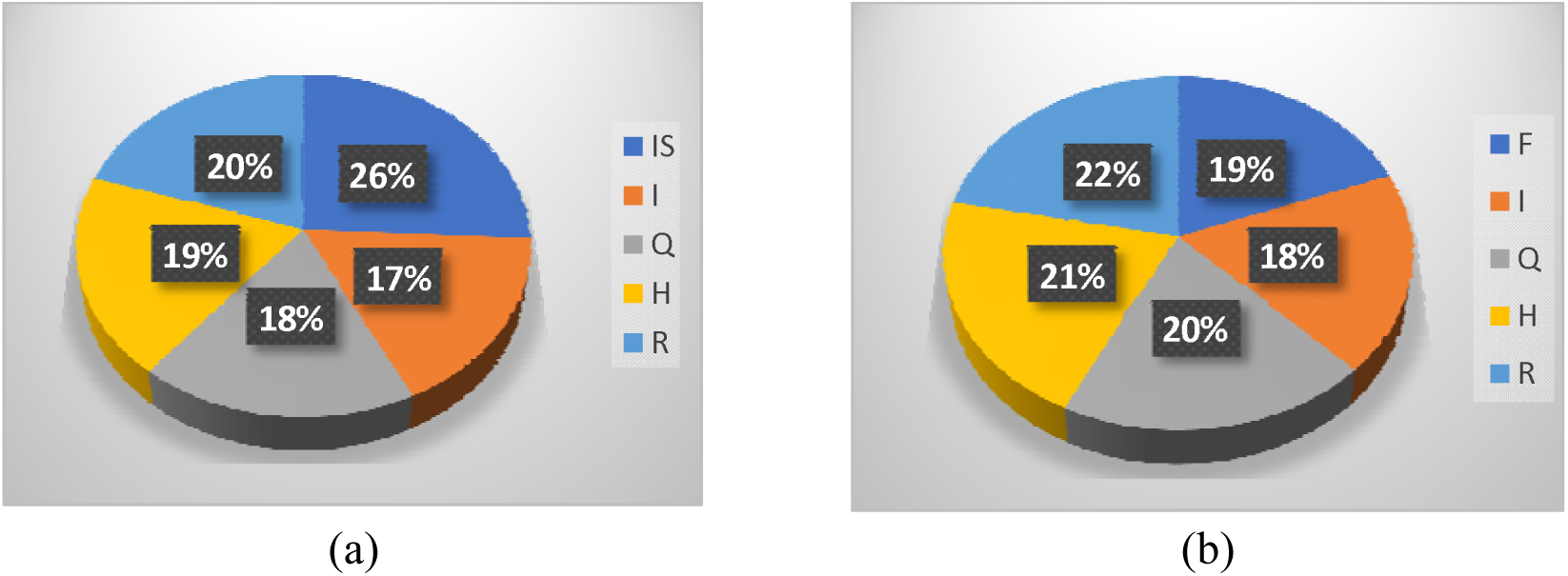
Percentage wise distribution of interstate and foreigner population in Covid-19 scenario

From the fig. 15 it can be observed that 7% of population is exposed to Covid-19. Interstate population share the largest percentage. 14% of the population is quarantined including foreigners and interstate population. Similarly, the infection is 13%. The hospitalization is done at 15%. Of the total population recovery is 16%.

**Fig. 15.**
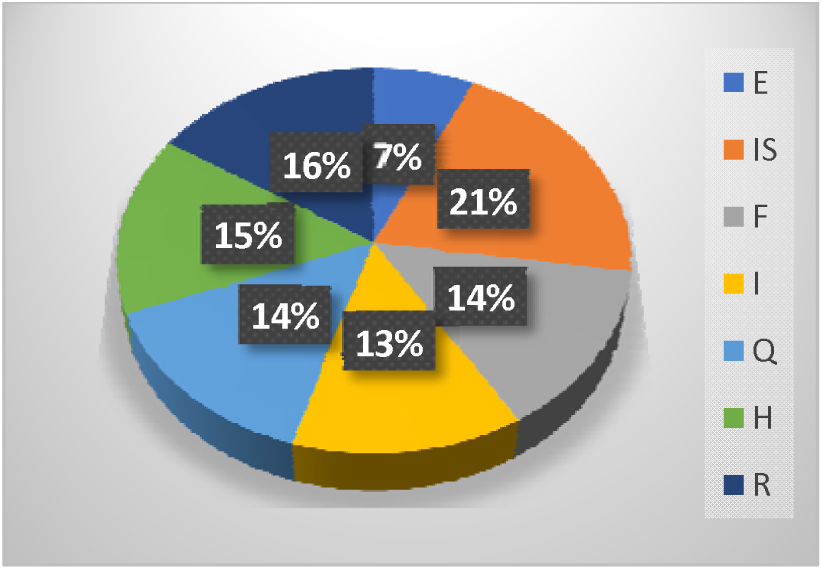
Percentage wise distribution of all the compartments of the model

## Discussion

Our model indicates that foreigners exhibit a larger infected population, hospitalization rate, and infection rate when compared to the interstate population. Moreover, as foreign individuals make contact with interstate individuals, the rate of infection within the interstate population increases significantly. To the best of our knowledge, our study is the first to create a disjoint mutually exclusive compartmental model. Our model suggests that both foreign and interstate travel lead to increased risk of infection within the United States population. Consequently, we validate the effectiveness of quarantine as a public health intervention model by the US government and encourage implementation of efforts to mitigate interstate travel.

There are multiple reasons that foreigners have increased risk of obtaining Covid-19 when compared to interstate population. First, as people travel, they risk exposing themselves to a greater number of other individuals. The WHO indicates that transmission of Covid-19 occurs primarily through droplet transmission (2019). Most methods of international travel, including airways, railways, and waterways, crowd individuals in compact and enclosed spaces. Being in close contact with individuals with respiratory symptoms in an enclosed environment increases the risk of being exposed to infected mucosae (Tatem et al. 2006). Second, the guidelines and strategies for addressing the epidemic differ among countries. For example, while India has enforced total lockdown, the US government have not mandated enforced lockdown (Chatterjee et al. 2020). Consequently, when individuals from countries with different regulations arrive, they may be infected and increase the incidence of covid-19. Finally, the vaccination standards differ among countries. In particular, BCG vaccine, believed to confer protective effects against Covid-19 is recommended in some countries, but not the US(Aaron Miller, Mac Josh Reandelar, Kimberly Fasciglione, Violeta Roumenova, Yan Li and Otazu 2013). As a result, future research and modelling is necessary to determine the protective effects of the BCG vaccine, and its potential to reduce the incidence of Covid-19 within the United States.

Given that 2019-nCoV is no longer contained within Wuhan, we recommend the United States government close their borders to both foreign and interstate travel. We recommend significant public health interventions at both international and interstate levels otherwise large cities with close inter-transport systems could become outbreak epicenters. Finally, we recommend preparedness plans and mitigation interventions be readied for quick deployment on both a state and federal level. Based on our model, compliance with these recommendations will effectively reduce the transmission of Covid-19 as a result of foreign and interstate travel.

## Data Availability

Our data derived from mathematical calculations done in the paper and is available to all the readers.

## Acknowledgement

The first three authors thank DST-FIST file # MSI-097 for technical support to the department. Third author (ENJ) is funded by UGC granted National Fellowship for Other Backward Classes (NFO-2018-19-OBC-GUJ-71790).

## Notes

### Competing Interest Statement

The authors have declared no competing interest.

### Funding Statement

The third author (ENJ) is funded by UGC granted National Fellowship for Other Backward Classes (NFO-2018-19-OBC-GUJ-71790).

### Author Declarations

This is not a clinical trial and no human is involved. IRB approval not required.

## Reference

Aaron Miller, Mac Josh Reandelar, Kimberly Fasciglione, Violeta Roumenova, Yan Li A, Otazu GH. Correlation between universal BCG vaccination policy and reduced morbidity and mortality for COVID-19: an epidemiological study Aaron. J Chem Inf Model. 2013;

Castillo-Chavez C, Castillo-Garsow CW, Yakubu AA. Mathematical Models of Isolation and Quarantine. Journal of the American Medical Association. 2003.

Chatterjee K, Chatterjee K, Kumar A, Shankar S. Healthcare impact of COVID-19 epidemic in India: A stochastic mathematical model. Med J Armed Forces India. 2020;

Cohen J, Normile D. New SARS-like virus in China triggers alarm. Science. 2020.

Diekmann O, Heesterbeek JAP, Roberts MG. The construction of next-generation matrices for compartmental epidemic models. J R Soc Interface. 2010;

Holshue ML, DeBolt C, Lindquist S, Lofy KH, Wiesman J, Bruce H, et al. First case of 2019 novel coronavirus in the United States. N Engl J Med. 2020;

Pontryagin LS. Mathematical Theory of Optimal Processes. Mathematical Theory of Optimal Processes. 2018.

Routh EJ. Routh, 1877. In: A treatise on the stability of a given state of motion: particularly steady motion, Macmillan Co.

Tang B, Wang X, Li Q, Bragazzi NL, Tang S, Xiao Y, et al. Estimation of the Transmission Risk of the 2019-nCoV and Its Implication for Public Health Interventions. J Clin Med. 2020;

Tatem AJ, Rogers DJ, Hay SI. Global Transport Networks and Infectious Disease Spread. Advances in Parasitology. 2006.

Wu JT, Leung K, Leung GM. Nowcasting and forecasting the potential domestic and international spread of the 2019-nCoV outbreak originating in Wuhan, China: a modelling study. Lancet. 2020;

Coronavirus disease (COVID-19) Pandemic [Internet]. World Health Organization. 2019 [cited 2020 Apr 14]. Available from: https://www.who.int/emergencies/diseases/novel-coronavirus-2019

Coronavirus (COVID-19) [Internet]. Centers for Disease Control and Prevention. 2020 [cited 2020 Apr 14]. Available from: https://www.cdc.gov/coronavirus/2019-ncov/index.html19

